# GutCore: An Endoscopy Foundation Model for Whole-Case Gastric Cancer Analysis

**DOI:** 10.64898/2026.07.01.26356993

**Authors:** Sehun Kim, Hyosoon Yoo, Seong-Keun Yoo, Jeeyun Lee, Hyuk Lee, Yang Won Min

## Abstract

**Background:** Routine endoscopy image sets contain complementary information about lesions, surrounding mucosa, anatomy, and examination context. However, most endoscopic artificial intelligence systems are developed and evaluated on selected frames for narrowly defined image-level tasks, limiting their ability to support case-level analysis.

**Objective:** To develop GutCore, an endoscopy foundation model for whole-case patient-level gastric cancer analysis, and assess whether routinely stored endoscopic image sets can support cancer detection, invasion-depth prediction, biomarker inference, and prognosis.

**Design:** GutCore was pretrained on 5.6 million de-identified endoscopic images from more than ten hospitals. We compared GutCore with general, medical, and endoscopy-specific foundation models using public image-level datasets and an internal retrospective cohort of 11,035 de-identified endoscopic examinations from Samsung Medical Center (2019–2023). Whole-case image sets were aggregated for patient-level prediction of cancer status, invasion depth, biomarker status, and overall survival.

**Results:** GutCore achieved AUCs of 0.996 for cancer detection, 0.960 for muscularis propria invasion, 0.801 for SM2-or-deeper invasion, and 0.781 for mucosal versus submucosal invasion among ESD-treated early gastric cancer cases. Biomarker prediction was strongest for EBV status and MLH1 loss and weaker for HER2 status, with AUCs of 0.861, 0.822, and 0.648, respectively. In the held-out advanced gastric cancer test set, GutCore-derived risk groups separated overall survival (log-rank *P* < .0001; high-risk vs low-risk hazard ratio, 13.18; 95% CI, 6.06–28.66), including within pathological stage II and III disease. Public image-level benchmarks further supported the generality of the learned endoscopic representation.

**Conclusions:** GutCore enabled whole-case patient-level gastric cancer assessment from routinely stored endoscopic images, extending endoscopy foundation model evaluation beyond selected frames. Independent external validation is required before clinical use.

## 1 Introduction

Gastrointestinal endoscopy is a routine first-line examination for the evaluation, surveillance, and treatment of digestive disease. During routine clinical practice, endoscopists capture and store images of major anatomical landmarks and suspected lesions to document examination quality, support diagnostic interpretation, and enable follow-up assessment. Because these images are acquired at scale during routine care, endoscopy is well suited to self-supervised visual pretraining. The unresolved question is whether such models can make patient-level predictions from whole endoscopic image sets, rather than only recognizing findings in curated single frames.

Many endoscopic artificial intelligence models have been developed for individual tasks, including colorectal polyp detection, gastric cancer detection, and prediction of gastric cancer invasion depth [1–4]. Although these models have shown promising performance, they are typically designed for a single clinical endpoint and are often evaluated using selected images with image-level classification or segmentation metrics. These evaluations capture frame-level performance but not how endoscopists actually work, integrating multiple images per case to reach a patient-level decision. Manual selection of representative images also requires expert review and is difficult to scale to large cohorts. This limitation is particularly relevant in gastric cancer because the stomach is a large and anatomically heterogeneous organ, and clinically relevant findings are not always captured in a single image. Assessment often requires integration of information across multiple views, including lesion morphology, extent, and the surrounding mucosal background. Moreover, some infiltrative gastric cancers, such as Borrmann type IV advanced gastric cancer, may show only subtle or diffuse endoscopic abnormalities without a well-demarcated visible mass, making diagnosis dependent on examination-wide contextual information rather than a single representative image. This motivates case-level evaluation in addition to image-level benchmarking.

Self-supervised vision models pretrained on large natural-image corpora have shown strong generalization across many visual tasks [5, 6]. In endoscopy, general vision foundation models have begun to be explored for tasks such as oropharyngeal cancer assessment, esophagogastric junction adenocarcinoma diagnosis, and gastrointestinal image retrieval [7–9]. These studies show that foundation models may be useful for endoscopic imaging, but endoscopy may still benefit from domain-specific pretraining because routine examinations generate correlated image sets with procedure-specific anatomy, image quality variation, and clinically relevant findings. Recent endoscopy-specific models, including Endo-FM, GastroNet-5M, and EndoMamba, have also shown that large-scale endoscopy pretraining is feasible for endoscopic image and video analysis [10–12]. Nevertheless, published evaluations of endoscopy foundation models remain concentrated on single-image classification and segmentation benchmarks, leaving open whether these models support patient-level endpoints.

In this study, we developed GutCore, a self-supervised vision foundation model for gastrointestinal endoscopy. GutCore was pretrained on approximately 5.6 million endoscopic images, anchored by GastroNet-5M, a 4.8-million-image multicenter collection from eight hospitals [11], and supplemented with private Samsung Medical Center (SMC) data, unlabeled HyperKvasir images [13], and captured frames from unlabeled HyperKvasir and GastroHUN endoscopy videos [14], yielding source data from more than ten hospitals overall. We evaluated GutCore against general and endoscopy-specific foundation models across both conventional image-level benchmarks and clinically oriented patient-level endpoints. At the image level, we assessed classification and segmentation performance on public endoscopy datasets. At the patient level, we aggregated whole-case endoscopic image sets for gastric cancer detection, depth prediction, biomarker prediction, and survival analysis among patients with advanced gastric cancer. The study therefore tested both curated frame-level performance and whole-case patient-level inference.

## 2 Methods

### 2.1 Study design

Figure 1 summarizes the whole-case analysis framework, evaluation scope, and GutCore pretraining strategy. In this retrospective study, we developed GutCore, an endoscopy foundation model, and evaluated it using internal patient-level gastric cancer cohorts and public image-level endoscopy datasets. Model selection was performed on validation splits, and final performance was reported on held-out test splits.

**Figure 1:**
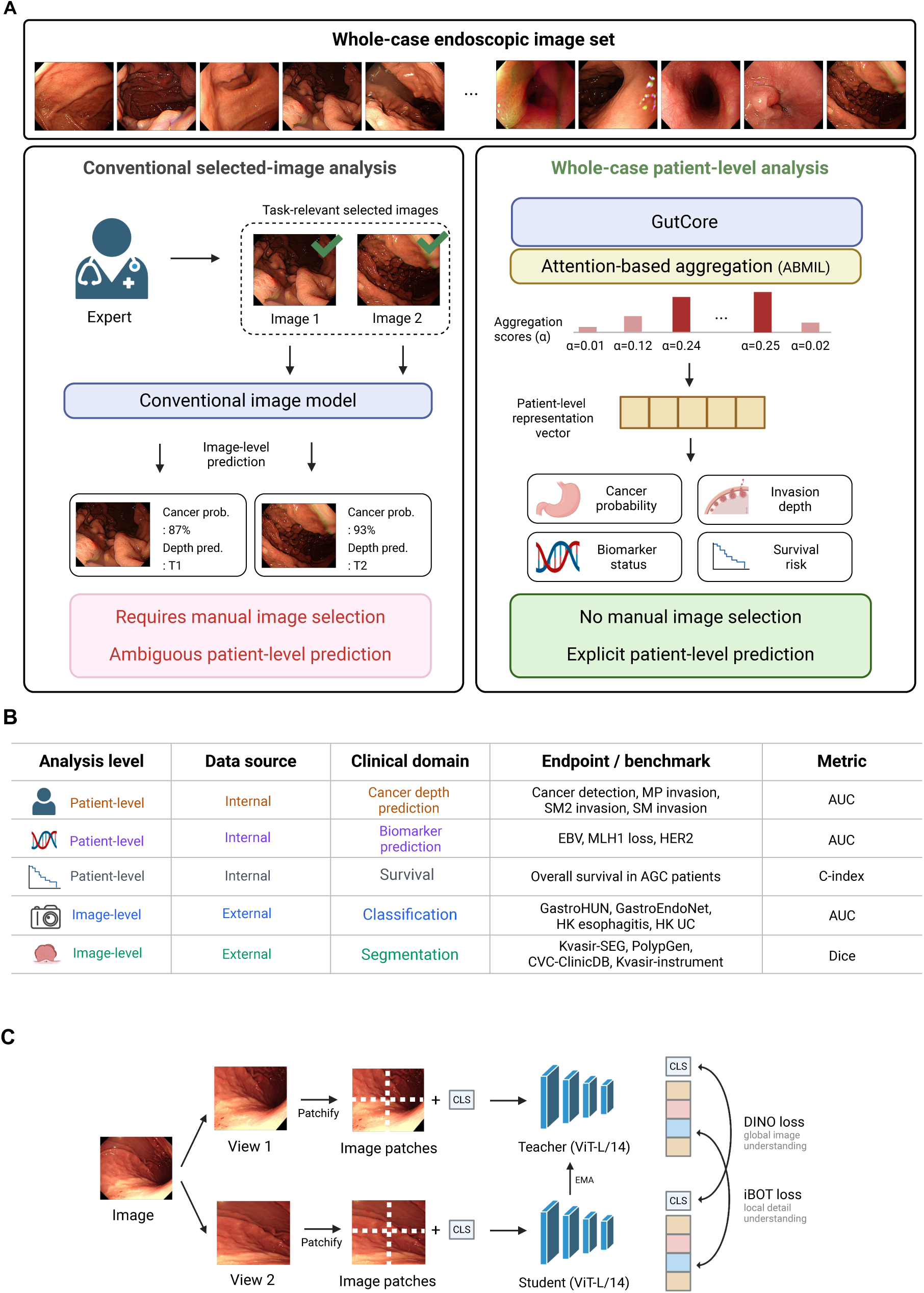
GutCore Whole-Case Analysis Framework and Study Overview. (A) Comparison of conventional selected-image analysis (left) and a whole-case framework (right). In conventional analysis, an expert selects task-relevant images, and an image-level model generates separate predictions without a predefined method for patient-level aggregation. In the whole-case framework, the contribution of each image is estimated dynamically, and information from the entire examination is combined to generate a single patient-level prediction. (B) Internal patient-level analyses and external image-level benchmarks. (C) Teacher-student self-supervised pretraining; the CLS token summarizes global image information, whereas patch tokens preserve local details. ABMIL, attention-based multiple-instance learning; AGC, advanced gastric cancer; AUC, area under the curve; C-index, concordance index; EMA, exponential moving average; HK, HyperKvasir; MP, muscularis propria; SM, submucosal; UC, ulcerative colitis. Figure created with BioRender.com.

### 2.2 Training data

GutCore was pretrained on a mixture of de-identified gastrointestinal endoscopy images assembled from multiple institutions and data sources. The largest component was GastroNet-5M, which contains roughly 4.8 million unlabeled endoscopic images from approximately 500,000 procedures across eight Dutch hospitals [11]. The remaining component included private SMC endoscopy data collected under the study protocol, comprising roughly 600,000 images from roughly 22,000 cases, approximately 100,000 unlabeled HyperKvasir images [13], and approximately 130,000 captured frames from HyperKvasir videos and GastroHUN unlabeled videoendoscopes [14].

The pretraining mixture sampled 70% of images from the GastroNet-5M component and 30% from the pooled non-GastroNet component. Additional information on the pretraining data and preprocessing is provided in Supplementary Section S1.2.

### 2.3 Development of the endoscopy foundation model

GutCore was developed as a self-supervised vision foundation model for gastrointestinal endoscopy. The model used a vision transformer (ViT) image encoder [15] and was pretrained on the multi-institutional endoscopy image corpus before task-specific evaluation. During pretraining, the model learned from unlabeled endoscopic images without task-specific labels such as cancer depth, biomarker status, or survival outcome.

The self-supervised training strategy was adapted from DINOv3 [6]. In brief, the model was trained to learn visual features from different views of the same endoscopic image and from images with partially masked regions. The objective was to learn general endoscopic visual patterns, including anatomy, mucosal texture, and lesion appearance, rather than features tied to a single predefined endpoint.

In the main analysis, the pretrained GutCore was kept frozen and only a lightweight prediction head was trained for each diagnostic, biomarker, or prognostic endpoint. End-to-end fine-tuning, in which both the encoder and the prediction model were updated for each task, was performed as a secondary analysis and is reported in Supplementary Section S4.1. Detailed architecture, pretraining objective, augmentation, and optimization settings are provided in Supplementary Section S2.1.

### 2.4 Validation of patient-level analyses in internal gastric cancer cohorts

To represent each archived endoscopic examination as a whole, patient-level analyses used all stored images from each de-identified index endoscopy rather than manually selected representative frames. This approach preserved examination-wide information while avoiding the expert review, operator dependence, and annotation burden associated with manual image selection. We used internal SMC cases collected between 2019 and 2023. Clinical variables, treatment history, endoscopy reports, pathology reports, biomarker test results, and survival status were obtained from DARWIN-C, the Clinical Data Warehouse of Samsung Medical Center.

The internal whole-case cohort included 11,035 cases: 8,049 pathology-confirmed gastric cancer cases and 2,986 non-cancer cases with benign gastritis or intestinal metaplasia. Gastric cancer was confirmed by pathological examination of ESD or surgical resection specimens, and intestinal metaplasia was confirmed by biopsy. Table 1 summarizes the overall internal cohort, and Table 2 shows the cohort, outcome definition, and outcome distribution for each patient-level analysis. Cancer detection used the full internal cohort, whereas invasion-depth, biomarker, and survival analyses were restricted to cases with the required pathological, biomarker, or follow-up information. Cancer detection distinguished gastric cancer from non-cancer cases. Invasion-depth analyses were performed in pathology-confirmed cancer cases and included three clinically relevant thresholds: submucosal (SM) invasion (T1a vs T1b) among ESD-treated EGC cases, SM2 invasion (M/SM1 vs SM2/SM3) among EGC cases, and EGC versus AGC, corresponding to invasion into the muscularis propria or beyond (T2 or higher). For the SM invasion, the cohort was restricted to ESD-treated EGC cases to evaluate a clinically narrow setting in which mucosal and submucosal invasion can be difficult to distinguish before treatment. The SM1/SM2 boundary is a key factor in deciding whether ESD alone is appropriate or surgical resection should be considered [16–18]. Biomarker analyses predicted EBV status, HER2 status, and MLH1 loss using pathology assay results obtained within 90 days of the index endoscopy. Detailed label definitions are provided in Supplementary Section S1.4.

**Table 1:** Clinical characteristics of the internal cohort. Values are n (%) unless otherwise indicated.

| Characteristic | Non-cancer | EGC | AGC |
| --- | --- | --- | --- |
| Number of cases | 2,986 | 6,168 | 1,881 |
| Age, median (IQR), years | 64 (57-70) | 62 (54-70) | 62 (54-70) |
| Male sex | 2,114 (70.8) | 4,022 (65.2) | 1,250 (66.5) |
| Stored images per examination, median (IQR) | 30 (25-37) | 31 (27-36) | 29 (25-33) |
| Treatment |  |  |  |
| Endoscopic submucosal dissection | – | 2,853 (46.3) | – |
| Surgical resection | – | 3,315 (53.7) | 1,881 (100.0) |
| Tumor location |  |  |  |
| Upper third/cardia/fundus | – | 1,175 (19.0) | 603 (32.1) |
| Middle third | – | 2,787 (45.2) | 713 (37.9) |
| Lower third/antrum/angle | – | 1,796 (29.1) | 445 (23.7) |
| Multiple lesions | – | 197 (3.2) | 44 (2.3) |
| Whole stomach/diffuse involvement | – | 0 (0.0) | 4 (0.2) |
| Remnant/anastomosis | – | 213 (3.5) | 72 (3.8) |
| Biomarker availability |  |  |  |
| EBV status | – | 3,539 (57.4) | 1,861 (98.9) |
| HER2 status | – | 3,596 (58.3) | 1,774 (94.3) |
| MLH1 status | – | 2,072 (33.6) | 1,082 (57.5) |
| Survival follow-up |  |  |  |
| Included in AGC survival cohort | – | – | 1,873 (99.6) |
| Follow-up, median (IQR), months | – | – | 38.6 (25.6-58.9) |
| Deaths during follow-up | – | – | 441 (23.5) |

AGC, advanced gastric cancer; EGC, early gastric cancer; ESD, endoscopic submucosal dissection; IQR, interquartile range.

**Table 2:** Internal analytic cohorts and outcome definitions for whole-case patient-level analyses in gastric cancer. Each endoscopic examination was analyzed as one whole-case input using all available stored images from the examination.

| Clinical endpoint | Analytic cohort | Prediction target | Outcome distribution |
| --- | --- | --- | --- |
| Gastric cancer detection | Full internal cohort | Non-cancer vs Gastric cancer | Non-cancer 2,986<br>Cancer 8,049 |
| SM invasion | ESD-treated EGC | M vs SM | M 2,235<br>SM 613 |
| SM2 invasion | EGC | M/SM1 vs SM2/SM3 | M/SM1 4,696<br>SM2/SM3 1,451 |
| MP invasion | GC | EGC vs AGC | EGC 6,168<br>AGC 1,881 |
| EBV status | GC with biomarker data | Negative vs positive | Negative 5,073<br>Positive 327 |
| HER2 status | GC with biomarker data | Negative vs positive | Negative 5,076<br>Positive 294 |
| MLH1 loss | GC with biomarker data | Intact/partial loss vs Complete loss | Intact/partial loss 2,872<br>Complete loss 282 |
| Overall survival | AGC | Overall survival risk score | Death 441<br>Censored 1,432 |

AGC, advanced gastric cancer; EGC, early gastric cancer; GC, gastric cancer; M, mucosal; SM, submucosal.

Survival analysis was restricted to surgically resected AGC cases. Overall survival was defined as the interval from the index endoscopy to death from any cause, with patients alive at the data cutoff censored at their last documented follow-up. AJCC 8th edition pathological stage served as the postoperative clinicopathological reference [19]. Model-derived risk scores were evaluated for survival discrimination and risk stratification; detailed stage and risk-group definitions are provided in Supplementary Section S1.4. For all patient-level predictions, image-level features were extracted from all examination images and aggregated into one case-level representation using attention-based multiple-instance learning (ABMIL) [20].

### 2.5 Validation on external image-level datasets

For image-level analyses, labels and predictions were assigned to individual frames or pixels rather than whole examinations. These external-dataset evaluations tested GutCore on conventional endoscopic image-analysis problems beyond the internal cohorts.

The classification tasks included GastroHUN stomach anatomical landmark classification (22 classes), HyperKvasir esophagitis grading (2 classes: Los Angeles grade A vs grades B-D), HyperKvasir ulcerative colitis grading (3 classes: Mayo scores 1, 2, and 3), and GastroEndoNet gastroesophageal reflux disease classification (2 classes: GERD vs non-GERD) [13, 14, 21].

The segmentation tasks included three colorectal polyp segmentation datasets, Kvasir-SEG, PolypGen, and CVC-ClinicDB, and one endoscopic instrument segmentation dataset, Kvasir-Instrument [22–25]. For these tasks, the target was a frame-level binary segmentation mask for the polyp or instrument region.

### 2.6 Comparison models

GutCore was compared with general vision, medical vision-language, and endoscopy-specific foundation models across all evaluation tasks. The comparison set included DINOv3 (ViT-B), MedGemma 1.5, GastroNet-5M, Endo-FM, and EndoMamba [6, 10–12, 26]. DINOv3 served as a general vision foundation-model baseline, MedGemma 1.5 as a medical foundation-model baseline, and GastroNet-5M, Endo-FM, and EndoMamba as endoscopy foundation models. For MedGemma 1.5, only the image encoder was used because the present evaluations tested visual features rather than text-generation performance. All comparison models were evaluated using the same evaluation procedures as GutCore, including image-level classification and segmentation, patient-level gastric cancer detection and depth prediction, biomarker prediction, and survival analysis.

### 2.7 Statistical analysis

For each endpoint, all models were evaluated using the same training, validation, and test split; predefined splits were used for public datasets when available. Models were trained on the training set, selected using the validation set, and evaluated on the held-out test set. Performance estimates are reported with 95% confidence intervals. For binary AUCs, confidence intervals were derived from DeLong variance estimates [27]; for multiclass AUCs, Dice coefficients, and C-indices, they were estimated using 2,000 bootstrap resamples at the relevant evaluation level (image or patient). *P* values were calculated using paired DeLong tests for binary AUCs and paired bootstrap tests for the other metrics. Survival analyses used Kaplan–Meier methods, global log-rank tests, and Cox proportional-hazards models. Further statistical details and robustness analyses are provided in Supplementary Section S3.1.

## 3 Results

### 3.1 Overview of whole-case gastric cancer and image-level benchmark evaluations

We evaluated GutCore first on whole-case patient-level gastric cancer endpoints and then on external image-level endoscopy benchmarks. The primary clinical analyses tested whether all stored images from an endoscopic examination could be aggregated to assess patient-level cancer status, invasion depth, biomarker status, and overall survival. These analyses used internal SMC gastric cancer cohorts and generated one prediction per endoscopic case.

We also tested the same pretrained model on conventional image-level datasets, including anatomical landmark recognition, disease grading, and segmentation. For the main analyses, the pretrained encoder was kept fixed and only task-specific prediction heads were trained. The main fixed-encoder results are shown in Figure 2, with end-to-end fine-tuning results provided in Supplementary Figure S1.

**Figure 2:**
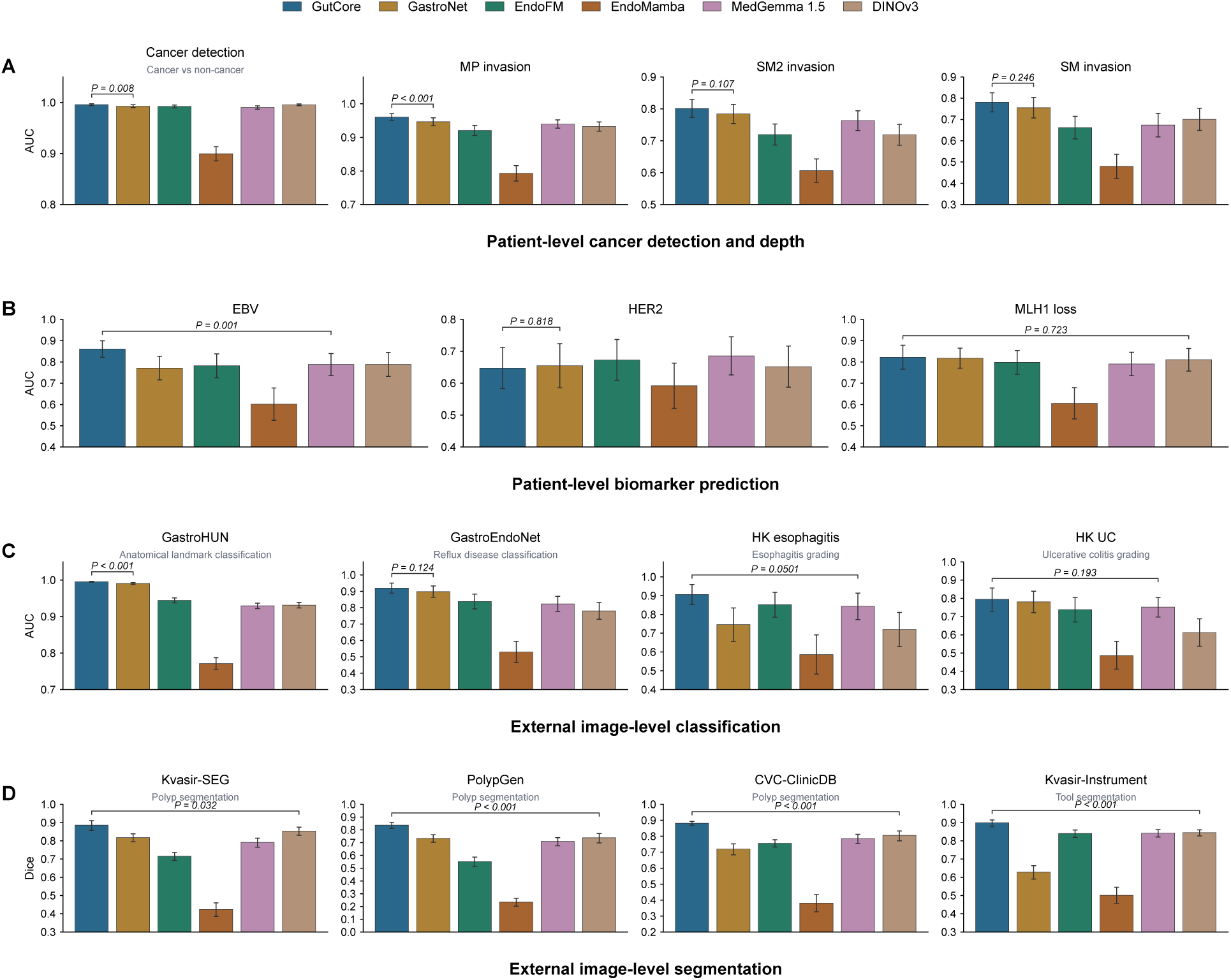
Whole-case patient-level gastric cancer analyses and image-level endoscopy benchmarks. (A,B) Patient-level cancer detection, depth assessment, and prediction of biomarker status in internal gastric cancer cohorts. (C,D) External image-level anatomical landmark recognition, disease grading, and segmentation benchmarks. Bars show held-out test-set estimates, and error bars show 95% confidence intervals (CIs) based on test images or patients. Bracketed *P* values compare GutCore with the non-GutCore model that had the highest validation-set performance. Binary AUCs use paired DeLong tests; multiclass AUCs and Dice coefficients use two-sided paired bootstrap tests.

### 3.2 Whole-case endoscopy supports patient-level cancer detection and invasion depth assessment

Gastric cancer assessment requires integration of lesion morphology, lesion extent, and surrounding mucosal context across the examination. We therefore evaluated whether whole-case endoscopic image sets could be used for patient-level cancer detection and clinically relevant invasion depth thresholds. Each prediction was made once per endoscopic case after aggregating all available images from that examination.

GutCore achieved high discrimination for cancer detection (AUC, 0.996; 95% CI, 0.994–0.998) and muscularis propria (MP) invasion (AUC, 0.960; 95% CI, 0.950–0.971; Figure 2A). Performance was lower for finer submucosal thresholds, including SM2-or-deeper invasion (AUC, 0.801; 95% CI, 0.773–0.829) and mucosal versus submucosal invasion among ESD-treated early gastric cancer cases (AUC, 0.781; 95% CI, 0.736–0.825). GutCore exceeded the validation-selected comparison model for cancer detection (*P* = 0.008) and MP invasion (*P* < 0.001), whereas the differences for the two finer submucosal thresholds were not significant.

This pattern is clinically plausible because broad separation between early and advanced gastric cancer is usually more visually apparent than fine submucosal depth stratification. Whole-examination aggregation performed best for broad early-versus-advanced distinctions, whereas finer submucosal thresholds remained more challenging.

### 3.3 Endoscopic image sets capture biomarker-associated visual phenotypes

We next asked whether whole-case endoscopic image sets contained visual signals associated with biomarker status. GutCore showed stronger discrimination for EBV status (AUC, 0.861; 95% CI, 0.822–0.900) and MLH1 loss (AUC, 0.822; 95% CI, 0.766–0.878) than for HER2 status (AUC, 0.648; 95% CI, 0.583–0.712; Figure 2B). GutCore exceeded the validation-selected comparison model for EBV status (*P* = 0.001), whereas the MLH1-loss and HER2 comparisons were not significant.

This heterogeneity suggests that some biomarkers may have more consistent endoscopic phenotypes than others. These results support the presence of biomarker-associated endoscopic phenotypes, most clearly for EBV status and MLH1 loss, but do not imply that white-light endoscopy can replace tissue-based biomarker testing.

### 3.4 External benchmarks show general endoscopic image performance

To confirm that GutCore learned generalizable endoscopic visual features, we evaluated it on open image-level classification and segmentation benchmarks. These benchmarks included gastric anatomical landmark classification, reflux and inflammatory disease classification, polyp segmentation, and endoscopic instrument segmentation. Detailed dataset characteristics, target labels, and split definitions for each benchmark are provided in Supplementary Section S1.3.

GutCore had the highest test-set estimate in all eight external benchmarks. Differences from the validation-selected comparison model were significant for GastroHUN and all four segmentation datasets, but not for the other three classification datasets (Figure 2). These external evaluations support the generality of GutCore as an endoscopic image representation, complementing the primary whole-case gastric cancer analyses.

### 3.5 GutCore-derived risk scores stratify survival in advanced gastric cancer

We evaluated whether whole-case endoscopic image-derived risk scores were associated with overall survival at the time of diagnostic endoscopy, including within pTNM stage strata.

In the held-out test set, GutCore-derived risk scores achieved a C-index of 0.644 (95% CI, 0.562–0.723; Figure 3A), with no significant difference from the validation-selected comparison model (*P* = 0.888). Risk groups defined using predefined cutoffs separated overall survival in the held-out AGC test set (log-rank *P <* 0.0001; Figure 3B). Using the low-risk group as reference, the hazard ratio was 3.53 for the moderate-risk group (95% CI, 1.60–7.77) and 13.18 for the high-risk group (95% CI, 6.06–28.66).

**Figure 3:**
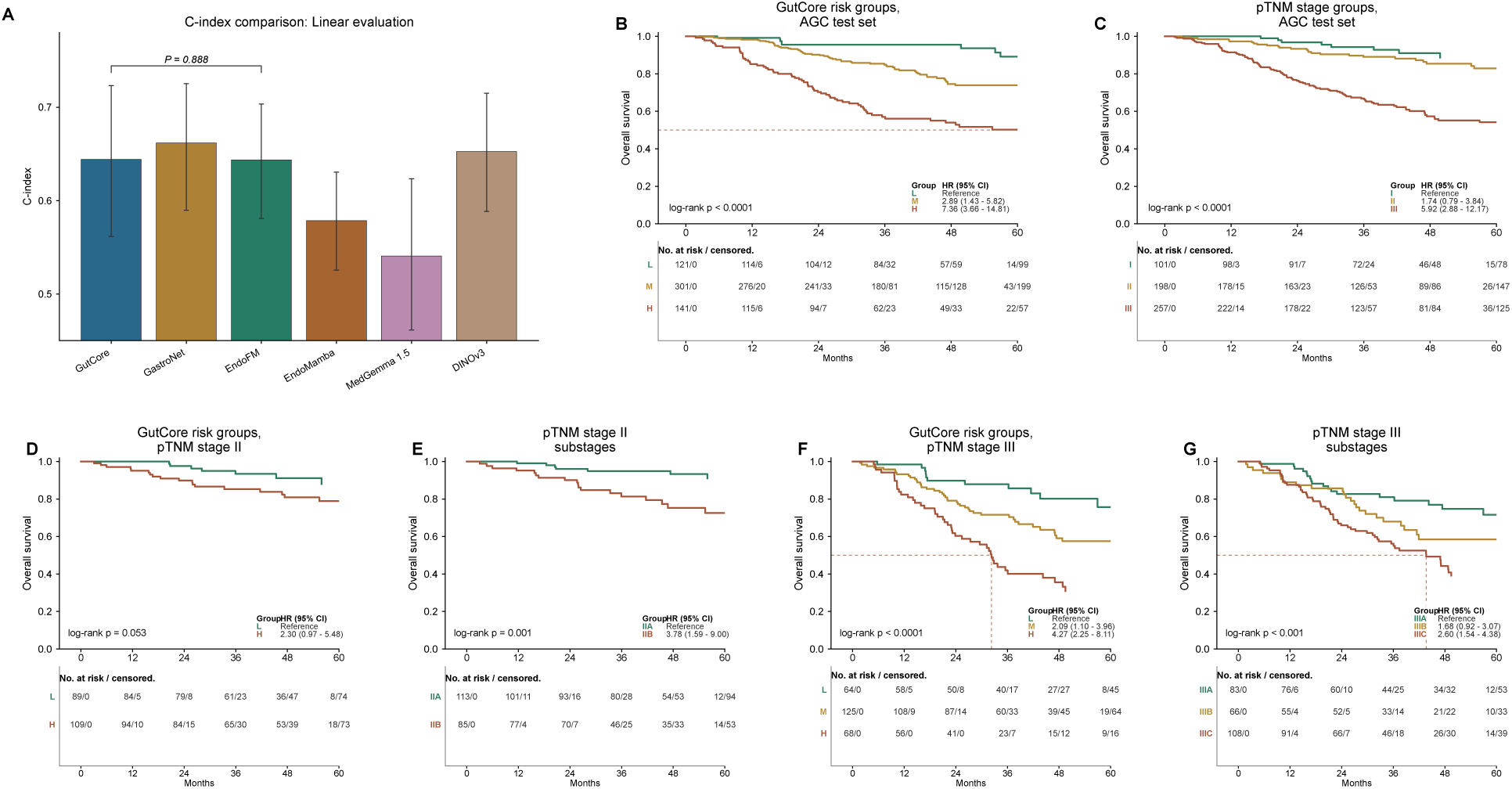
Survival analysis in advanced gastric cancer. (A) Test-set C-index with the pretrained encoder kept fixed; error bars show 95% CIs from paired test-patient bootstrap resampling. The bracketed *P* value compares GutCore with the non-GutCore model that had the highest validation-set C-index using a two-sided paired bootstrap test. (B,C) Kaplan–Meier curves for GutCore-derived risk groups and pTNM stage in the same AGC test set. (D–G) Survival stratification within pTNM stage II and stage III disease, showing GutCore-derived risk groups and corresponding pTNM substages. AGC, advanced gastric cancer; CI, confidence interval; pTNM, pathological tumor-node-metastasis.

For clinical context, Kaplan–Meier curves based on pTNM stage are shown in parallel (Figure 3C). GutCore-derived risk groups also separated survival within pTNM stage II and stage III disease (Figure 3D,F), with corresponding pTNM substage curves shown for comparison (Figure 3E,G).

These results indicate that whole-case endoscopic images contain prognostic information within pTNM stage strata, although the analysis remains exploratory.

### 3.6 Attention maps support qualitative interpretation of whole-case predictions

We performed attention analyses to review whole-case predictions at both the case-image and within-image levels. In the attention-based aggregation model, each image from an examination receives an aggregation weight; images with higher weights contribute more to the final case-level prediction. We then inspected [CLS] token attention maps within images to obtain a visual summary of the image regions emphasized by the model. These maps provide a way to check whether the images and regions emphasized by the model correspond to recognizable endoscopic abnormalities. Across representative cancer-detection and MP-invasion examples, high-weighted images generally contained lesion-related mucosal abnormalities, whereas low-weighted images were less visually informative for the case-level prediction (Figure 4). Spatial attention maps emphasized corresponding abnormal regions within selected high-weighted images.

**Figure 4:**
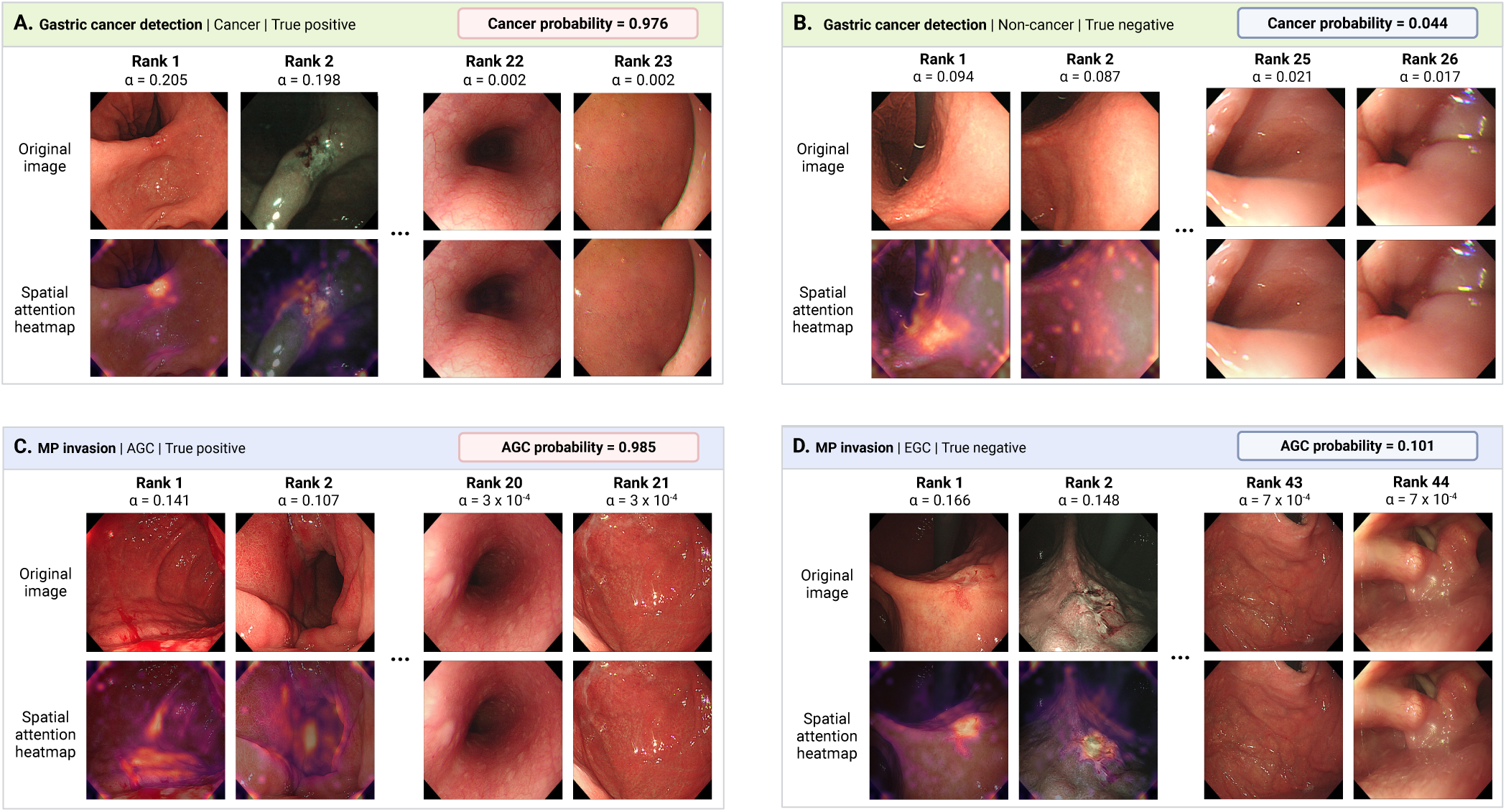
Qualitative attention analysis for whole-case classification. (A,B) Gastric cancer detection examples from cancer and non-cancer cases. (C,D) MP-invasion examples from Borrmann type IV advanced gastric cancer and early gastric cancer cases. Aggregation scores (*α*) rank images within each case, and [CLS] token attention heatmaps indicate spatially emphasized regions within selected images. AGC, advanced gastric cancer; EGC, early gastric cancer; MP, muscularis propria. Figure created with BioRender.com.

The cancer-detection example showed concentrated aggregation weights in lesion-containing images, whereas the non-cancer example showed a more distributed pattern, consistent with the absence of a focal malignant finding. In the MP-invasion example, the true-positive AGC case showed Borrmann type IV morphology, for which endoscopic abnormalities may be diffuse rather than mass-forming; high-weighted images and spatial maps emphasized wall-thickening and fold-related abnormalities compatible with this phenotype. These analyses are qualitative and should be interpreted as case-review examples rather than validated causal explanations.

### 3.7 Embedding visualization and retrieval illustrate clinically recognizable gastric cancer phenotypes

We examined whether GutCore image embeddings reflected clinically recognizable gastric cancer phenotypes. UMAP [28] was used as a descriptive two-dimensional visualization of the image-level embedding space, and query-centered local neighborhoods were inspected around representative EGC and AGC query images (Figure 5A–C). For the EGC query, retrieved images showed similar subtle flat or depressed lesions and early pathological stage; one retrieved case treated by ESD lacked nodal staging (Figure 5D). For the AGC query, retrieved cases shared Borrmann type IV gross morphology, diffuse endoscopic features, and poorly cohesive or signet-ring-cell components (Figure 5E). These examples suggest that GutCore embeddings can retrieve visually and clinically similar cases, including infiltrative phenotypes that may be difficult to summarize with a single local image feature. This analysis was qualitative and should not be interpreted as a validated clustering analysis or diagnostic retrieval benchmark.

**Figure 5:**
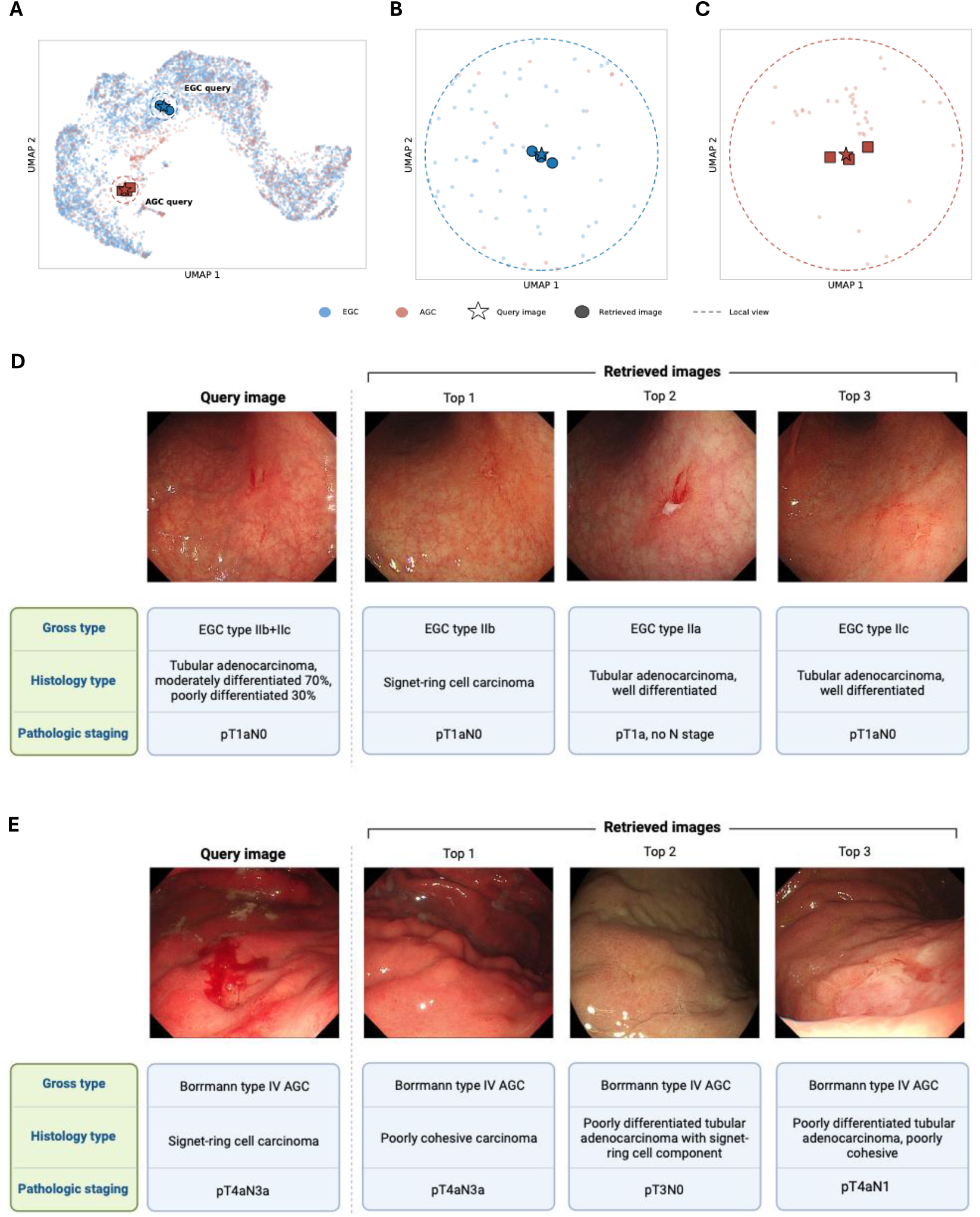
GutCore embedding space and image-retrieval examples. (A) UMAP visualization of image-level GutCore embeddings from EGC and AGC images. Stars indicate query images, filled markers indicate retrieved images, and point colors indicate EGC or AGC. (B,C) Query-centered local embedding neighborhoods for the EGC query and Borrmann type IV AGC query, respectively. Dashed circles indicate the radius used to define the displayed local view. (D) EGC query image and top-three retrieved images with corresponding gross type, histology, and pathological stage. (E) AGC query image and top-three retrieved images, showing similar diffuse morphology and poorly cohesive or signet-ring-cell-related histology. AGC, advanced gastric cancer; EGC, early gastric cancer; ESD, endoscopic submucosal dissection; UMAP, uniform manifold approximation and projection. Figure created with BioRender.com.

## 4 Discussion

This study developed GutCore and evaluated whether whole-case endoscopic image sets could support patient-level gastric cancer assessment beyond selected single-frame analysis. The principal finding was that aggregating all stored images from an endoscopic examination supported cancer detection, evaluation of clinically relevant invasion depth, biomarker prediction, and prognostic assessment. External image-level benchmarks further supported the generality of GutCore across diverse endoscopic imaging tasks, but the main clinical contribution is the extension from curated frame-level evaluation to whole-case patient-level inference.

Whole-case analysis offers several clinical and practical advantages. It reflects the review of multiple views from an endoscopic examination rather than relying on one representative image. It also avoids endpoint-specific frame selection, which requires expert effort and may need to be repeated because the most informative images can differ across clinical questions. By generating one prediction per examination, whole-case analysis aligns the unit of analysis with the patient-level clinical endpoint. This is particularly relevant to gastric cancer, for which lesion morphology, extent, background mucosa, and gastric anatomy may all contribute to the final impression. GutCore showed high discrimination for cancer detection and muscularis propria invasion, whereas performance was lower for finer submucosal thresholds.

The biomarker and prognostic findings were particularly noteworthy because these endpoints extend beyond the conventional visual assessment of gastric cancer during endoscopy. EBV status and MLH1 loss showed stronger image-associated signals than HER2 status, suggesting that some biomarker states may be associated with more consistent endoscopic phenotypes than others. These associations may reflect tumor location, gross morphology, histologic phenotype, background mucosal changes, or other clinicopathologic correlates. Similarly, GutCore-derived risk groups separated overall survival and identified prognostically distinct subgroups within pTNM stage II and III disease.

The qualitative attention and retrieval analyses provided examples of how the learned representation could be inspected. Image-level aggregation scores identified which images contributed most to case-level predictions, and spatial attention maps highlighted regions emphasized within selected images. Retrieval examples illustrated the potential for similar-case review by retrieving images with endoscopic features similar to those of the query case.

Several limitations should be considered. The patient-level analyses were retrospective and based on internal clinical cohorts, so external validation in locked independent cohorts is required. Although pretraining used multi-institutional data, this does not constitute external clinical validation of the gastric cancer endpoints. Patient-level labels were derived from routine clinical, pathological, and biomarker records, and the biomarker and survival analyses may therefore be influenced by biomarker testing patterns and cohort selection.

## Acknowledgement

This research was supported by a grant of the Korea Health Technology R&D Project through the Korea Health Industry Development Institute (KHIDI), funded by the Ministry of Health & Welfare, Republic of Korea (grant number: RS-2024-00437038). This research was also supported by the Bio&Medical Technology Development Program of the National Research Foundation (NRF), funded by the Korean government (MSIT) (No. RS-2023-00222838).

## 5 Data Availability

Code, model resources, and project information are available at https://github.com/SMC-GutX/ GutCore. The public image-level datasets used in this study are available from their original sources, as cited in the Methods and Supplementary Section S1.3. The internal Samsung Medical Center data is not publicly available.

## Supplementary Material

### S1. Data Sources and Cohorts

#### S1.1. Ethics and Privacy

Private Samsung Medical Center (SMC) data were handled as de-identified retrospective endoscopy data under institutional data-governance procedures. Use of the internal retrospective SMC data was approved by the Institutional Review Board at SMC (No. 2025-09-081). The requirement for informed consent was waived by the Institutional Review Board because of the retrospective study design. Public datasets were used according to their data-use terms and availability.

#### S1.2. Pretraining Data and Preprocessing

The pretraining data loader used a weighted image-folder mixture. Images were sampled 70% from GastroNet-5M and 30% from pooled non-GastroNet data sources. GastroNet-5M contributed 4.8 million unlabeled images from approximately 500,000 procedures across eight hospitals. The non-GastroNet component added private Samsung Medical Center (SMC) data and public unlabeled data to the pretraining distribution. The private SMC component contained roughly 600,000 images from roughly 22,000 cases. The public component added approximately 100,000 unlabeled HyperKvasir images and 130,000 frames captured from HyperKvasir and GastroHUN endoscopy videos [13, 14]. Images were normalized using channel statistics estimated from pretraining images, with mean [0.571, 0.329, 0.252] and standard deviation [0.274, 0.198, 0.171].

#### S1.3. External Image-Level Benchmark Datasets

External image-level benchmarks were used to test general endoscopic visual recognition and segmentation. The classification benchmarks evaluated frame-level recognition across anatomical, inflammatory, and reflux-related labels, whereas the segmentation benchmarks evaluated lesion or tool localization at the image-mask level (Table S1). For CVC-ClinicDB and Kvasir-Instrument, stratified 70/10/20 train/validation/test partitions were generated for independent experiments. For datasets with predefined or previously fixed split files, the same split assignment was preserved across independent experiments.

#### S1.4. Internal Patient-Level Datasets

The internal patient-level cohort was assembled from SMC endoscopy cases between 2019 and 2023 whose final pathology established early gastric cancer (EGC) or advanced gastric cancer (AGC). Eligible EGC cases were those treated by endoscopic submucosal dissection (ESD) or surgical resection, whereas eligible AGC cases were those treated by surgical resection. Accordingly, EGC or AGC patients without these qualifying procedures were not included in the current internal patient-level evaluation, including AGC patients whose clinical status was too poor to undergo surgery. The T1a/T1b task distinguished T1a mucosal cancer (M) from T1b submucosal cancer (SM) among ESD-treated EGC cases. The SM2 invasion task was defined among EGC cases treated by ESD or surgical resection and classified whether invasion depth was no deeper than SM1 or extended to SM2 or beyond. In this task, M and SM1 were grouped as the shallower class, whereas SM2 and SM3 were grouped as the deeper class. Although SM1, SM2, and SM3 are all subdepths within T1b submucosal invasion, the SM1/SM2 boundary is clinically important because it is a major factor in deciding between ESD alone and surgical resection with lymph node assessment. The muscularis propria (MP) invasion task defined the broad internal cohort by assigning EGC cases to T1 disease and AGC cases to T2 or deeper disease. After requiring matched endoscopy, treatment, and pathology records, at least eight endoscopic images per case, and concordance between the EGC/AGC source label and extracted pathological T category, the MP invasion task contained 8,050 cases, including 6,168 EGC cases and 1,882 AGC cases. The train, validation, and test splits contained 4,896, 1,633, and 1,633 cases.

**Table S1:** External image-level benchmark datasets. Split counts are reported as train/validation/test. For CVC-ClinicDB and Kvasir-Instrument, the counts shown correspond to the split used for the main analysis; new stratified random splits were generated for each independent experiment.

| Dataset | Endpoint | Split | Split protocol |
| --- | --- | --- | --- |
| GastroHUN [14] | Stomach anatomical landmark recognition; 22 classes | 3,641/777/784 | Fixed split |
| HK esophagitis [13] | Esophagitis grading; LA grade A vs grades B–D | 397/133/133 | Random split |
| HK UC [13] | Ulcerative colitis grading; Mayo scores 1–3 | 466/155/156 | Random split |
| GastroEndoNet [21] | GERD detection; GERD vs non-GERD | 1,557/208/312 | Random split |
| Kvasir-SEG [22] | Colon polyp segmentation | 600/200/200 | Random split |
| PolypGen [23] | Colon polyp segmentation | 1,013/227/296 | Fixed split |
| CVC-ClinicDB [25] | Colon polyp segmentation | 428/61/123 | Random split |
| Kvasir-Instrument [24] | GI endoscopic tool segmentation | 413/59/118 | Random split |

The survival cohort included 1,873 surgically resected AGC cases without documented distant metastasis, among which 441 deaths were observed and 1,432 cases were censored. Overall survival was defined as the interval from the index endoscopy to death from any cause; patients alive at the data cutoff were censored at their last documented follow-up. Postoperative stage grouping followed the AJCC 8th edition gastric cancer pTNM system [19]. The full stage map is shown in Table S2; stage grouping in this non-metastatic cohort was based on pathological tumor depth and nodal status within M0 disease. Stage I in Figure 3 combines AJCC stage IA and IB, stage II combines IIA and IIB, and stage III combines IIIA, IIIB, and IIIC.

**Table S2:**
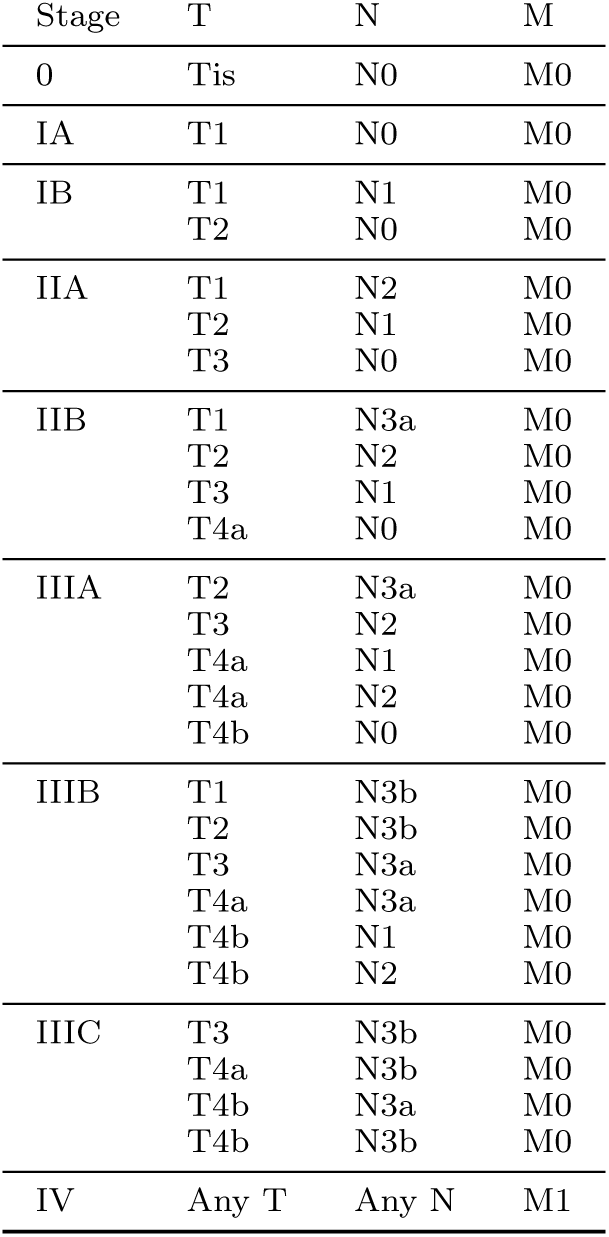
AJCC 8th edition pTNM stage groups for gastric cancer. The survival analyses used the applicable surgically resected non-metastatic stage groups from this stage map.

| Stage | T | N | M |
| --- | --- | --- | --- |
| 0 | Tis | N0 | M0 |
| IA | T1 | N0 | M0 |
| IB | T1 | N1 | M0 |
|  | T2 | N0 | M0 |
| IIA | T1 | N2 | M0 |
|  | T2 | N1 | M0 |
|  | T3 | N0 | M0 |
| IIB | T1 | N3a | M0 |
|  | T2 | N2 | M0 |
|  | T3 | N1 | M0 |
|  | T4a | N0 | M0 |
| IIIA | T2 | N3a | M0 |
|  | T3 | N2 | M0 |
|  | T4a | N1 | M0 |
|  | T4a | N2 | M0 |
|  | T4b | N0 | M0 |
| IIIB | T1 | N3b | M0 |
|  | T2 | N3b | M0 |
|  | T3 | N3a | M0 |
|  | T4a | N3a | M0 |
|  | T4b | N1 | M0 |
|  | T4b | N2 | M0 |
| IIIC | T3 | N3b | M0 |
|  | T4a | N3b | M0 |
|  | T4b | N3a | M0 |
|  | T4b | N3b | M0 |
| IV | Any T | Any N | M1 |

Ground-truth biomarker labels were obtained from clinical pathology assays performed within 90 days of the index endoscopy. The final biomarker cohorts after excluding records with incomplete or inconsistent pathology information were EBV status, 5,400 cases with 327 positive; HER2 status, 5,370 cases with 294 positive; and MLH1 loss, 3,154 cases with 282 positive. EBV status was assessed by in situ hybridization for EBV-encoded RNA (EBER-ISH), with positive and negative reports mapped to the corresponding binary labels. HER2 was assessed by IHC using an anti-HER2 monoclonal antibody (clone 4B5) on the Ventana BenchMark ULTRA platform (Roche Diagnostics). An IHC score of 3+ was classified as positive, scores of 0 or 1+ as negative, and equivocal 2+ results as unavailable for the binary HER2 analysis. MLH1 status was assessed by IHC; complete loss was classified as MLH1 loss, whereas intact, preserved, non-complete, or partial loss patterns were classified as no complete loss. Prepared biomarker splits used a 70/15/15 train, validation, and test ratio with stratification.

Model-derived risk scores were evaluated using the inverse-probability-of-censoring weighted C-index. Survival analyses were conducted using Kaplan–Meier methods and Cox proportional-hazards models. Survival curves were compared using global log-rank tests, and Cox models were used to estimate hazard ratios with 95% confidence intervals. Following a previously used stratification approach [29], the overall AGC and stage III analyses defined low-, moderate-, and high-risk groups using the first and third quartiles of the corresponding risk-score distributions in the training and validation data. The stage II analysis defined low- and high-risk groups using the median risk score among stage II training and validation cases. These thresholds were then applied without modification to the corresponding held-out test patients.

### S2. Model Architecture and Whole-Case Modeling

#### S2.1. GutCore Architecture and Self-Supervised Training

The primary GutCore configuration used a ViT-L/14 vision transformer initialized without pretrained weights and without register or storage tokens. Pretraining combined a DINO-style global-view objective and an iBOT-style masked-patch objective using the DINOv3 training engine, with additional KDE entropy regularization to encourage feature diversity. Teacher-student centering used Sinkhorn-Knopp centering, and the teacher model was updated by exponential moving average. The complete pretraining configuration is summarized in Table S3.

**Table S3:**
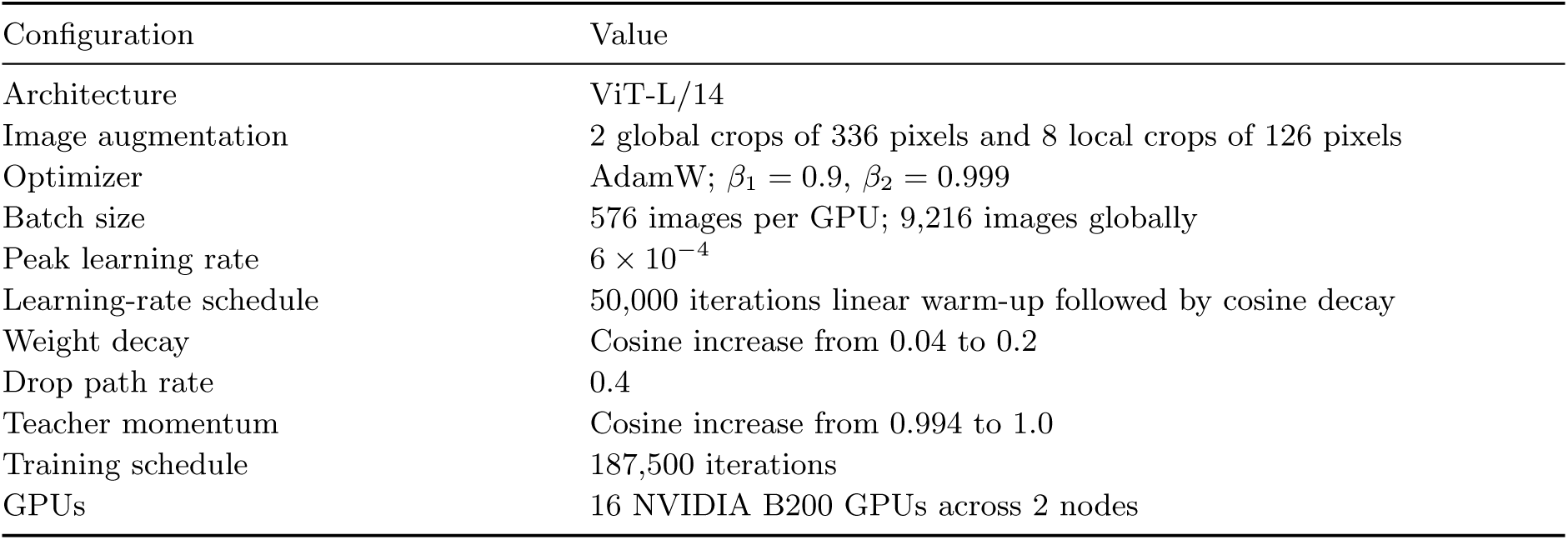
GutCore pretraining configuration.

| Configuration | Value |
| --- | --- |
| Architecture | ViT-L/14 |
| Image augmentation | 2 global crops of 336 pixels and 8 local crops of 126 pixels |
| Optimizer | AdamW; $\beta_1 = 0.9$, $\beta_2 = 0.999$ |
| Batch size | 576 images per GPU; 9,216 images globally |
| Peak learning rate | $6 \times 10^{-4}$ |
| Learning-rate schedule | 50,000 iterations linear warm-up followed by cosine decay |
| Weight decay | Cosine increase from 0.04 to 0.2 |
| Drop path rate | 0.4 |
| Teacher momentum | Cosine increase from 0.994 to 1.0 |
| Training schedule | 187,500 iterations |
| GPUs | 16 NVIDIA B200 GPUs across 2 nodes |

#### S2.2. Attention-Based Multiple-Instance Learning for Whole-Case Prediction

Whole-case gastric cancer endpoints were modeled with attention-based multiple-instance learning (ABMIL), because each endoscopic examination contained a variable number of stored images but only one case-level label. For a case *i*, all available images were treated as a bag *B_i_* = {*x_i_*_1_*, . . . , x_in__i_* }. The image encoder produced one feature vector *h_ij_*for each image. In frozen-encoder analyses, these image features were precomputed and kept fixed; in fine-tuning analyses, the image encoder, ABMIL module, and prediction head were updated together.

The ABMIL module used gated attention [20]. After layer normalization, each image feature was passed through two attention branches, and the attention score was computed as

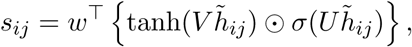

where *h̃_ij_* denotes the normalized image feature, ⊙ denotes element-wise multiplication, and *σ* denotes the sigmoid function. Scores were converted into image-level aggregation weights by a softmax over all images in the case,

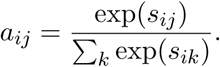

The case representation was the weighted sum *z_i_* = Σ_j_*a_ij_h̃_ij_*. A linear classification head mapped *z_i_* to class logits for cancer detection, invasion-depth assessment, and biomarker status. For survival analysis, the same gated-attention pooling produced a case representation that was mapped by a linear Cox risk head to a scalar risk score.

During patient-level classification training, up to 8 images were randomly sampled from each case at each training step to reduce computation and provide stochastic regularization. Validation and test evaluation used all available images from each case. The aggregation weights were also used for qualitative case review, where high-weighted and low-weighted images were inspected together with within-image attention maps.

#### S2.3. Implementation details for evaluation analyses

Linear evaluation was used to assess pretrained features with the encoder kept fixed, and fine-tuning was used to assess task-specific adaptation after self-supervised pretraining. All linear-evaluation experiments used the same train, validation, and test splits as the corresponding fine-tuning experiments. Fine-tuning used validation-based model selection and early stopping. Image classification used AdamW optimization for GutCore and DINOv3, with model-specific optimization recipes for other baselines. Segmentation used AdamW with layer-wise learning-rate decay for the default recipe and model-specific loss or scheduler variants for some baselines. Case-level and biomarker fine-tuning used weighted sampling for imbalanced labels, warm-up, gradient clipping, label smoothing, and validation area under the receiver operating characteristic curve for checkpoint selection. During case-level training, up to 8 images were randomly sampled from each case to reduce computation and regularize multiple-instance learning; evaluation used all available images per case.

### S3. Evaluation and Qualitative Analyses

#### S3.1. Statistical and Evaluation Details

Classification endpoints were summarized using area under the receiver operating characteristic curve (AUC), segmentation endpoints using Dice coefficient, and survival discrimination using the inverse-probability-of-censoring weighted concordance index (C-index). Survival analyses were conducted using Kaplan–Meier methods and Cox proportional-hazards models. Survival curves were compared using global log-rank tests, and Cox models were used to estimate hazard ratios with 95% confidence intervals. Model training, evaluation, and statistical analyses were performed in Python using PyTorch, scikit-learn, SciPy, and scikit-survival.

##### Primary test-set estimates and confidence intervals

The main figures and numerical results use a fixed training, validation, and test split for each endpoint. All models were evaluated on the same test observations, and public datasets with predefined partitions retained their original test sets. Binary AUCs and 95% confidence intervals (CIs) were calculated using DeLong’s method. Multiclass macro-AUC CIs were estimated using 2,000 class-stratified test-image bootstrap resamples. Dice CIs used 2,000 paired test-image bootstrap resamples, with each image treated as the resampling unit. Inverse-probability-of-censoring weighted C-index CIs used 2,000 paired test-patient bootstrap resamples while retaining the censoring distribution estimated from the training set. Hazard-ratio CIs in the Kaplan–Meier panels were estimated from Cox proportional-hazards models in the same held-out test split.

##### Model comparisons

For each endpoint, GutCore was compared with the non-GutCore model that had the highest performance on the validation set. Binary AUC differences were tested using two-sided paired DeLong tests. Multiclass AUC, Dice, and C-index differences were tested using two-sided paired bootstrap tests with the same resampled test observations used for both models. Reported *P* values were interpreted descriptively as task-level comparisons and were not adjusted for multiplicity.

##### Robustness across data splits and independent experiments

Fixed-encoder classification and segmentation were evaluated in 10 independent experiments, and fixed-encoder survival analysis in 20 (Supplementary Table S4). These analyses incorporated variation in data partitioning and training stochasticity but were not used to construct the primary test-sample CIs in Figures 2 and 3. Because the data partitions overlapped, experiment-level estimates were not treated as independent test samples.

**Table S4:** Robustness of GutCore performance across independent experiments. Values are mean test performance (95% CI) across experiments, calculated using Student’s *t* distribution. These intervals summarize variation across data partitions and independent experiments, not uncertainty from resampling the fixed test set.

| Endpoint | Metric | Experiments | Mean (95% CI) |
| --- | --- | --- | --- |
| Cancer detection | AUC | 10 | 0.995 (0.995–0.996) |
| MP invasion | AUC | 10 | 0.960 (0.956–0.963) |
| SM2 invasion | AUC | 10 | 0.804 (0.798–0.810) |
| SM invasion | AUC | 10 | 0.759 (0.741–0.776) |
| EBV | AUC | 10 | 0.831 (0.811–0.851) |
| HER2 | AUC | 10 | 0.673 (0.649–0.698) |
| MLH1 loss | AUC | 10 | 0.854 (0.838–0.870) |
| GastroHUN | AUC | 10 | 0.996 (0.996–0.996) |
| GastroEndoNet | AUC | 10 | 0.918 (0.914–0.922) |
| HK esophagitis | AUC | 10 | 0.883 (0.867–0.900) |
| HK UC | AUC | 10 | 0.776 (0.758–0.793) |
| Kvasir-SEG | Dice | 10 | 0.885 (0.884–0.886) |
| PolypGen | Dice | 10 | 0.839 (0.836–0.842) |
| CVC-ClinicDB | Dice | 10 | 0.883 (0.879–0.887) |
| Kvasir-Instrument | Dice | 10 | 0.902 (0.899–0.905) |
| AGC survival | C-index | 20 | 0.670 (0.652–0.688) |

### S4. Supporting Experimental Results

#### S4.1. End-to-End Fine-Tuning Performance

End-to-end fine-tuning was evaluated on the same tasks and comparison models as the frozen-feature evaluation. This analysis tested whether updating the encoder and task head changed the performance pattern observed with the encoder kept fixed.

**Figure S1:**
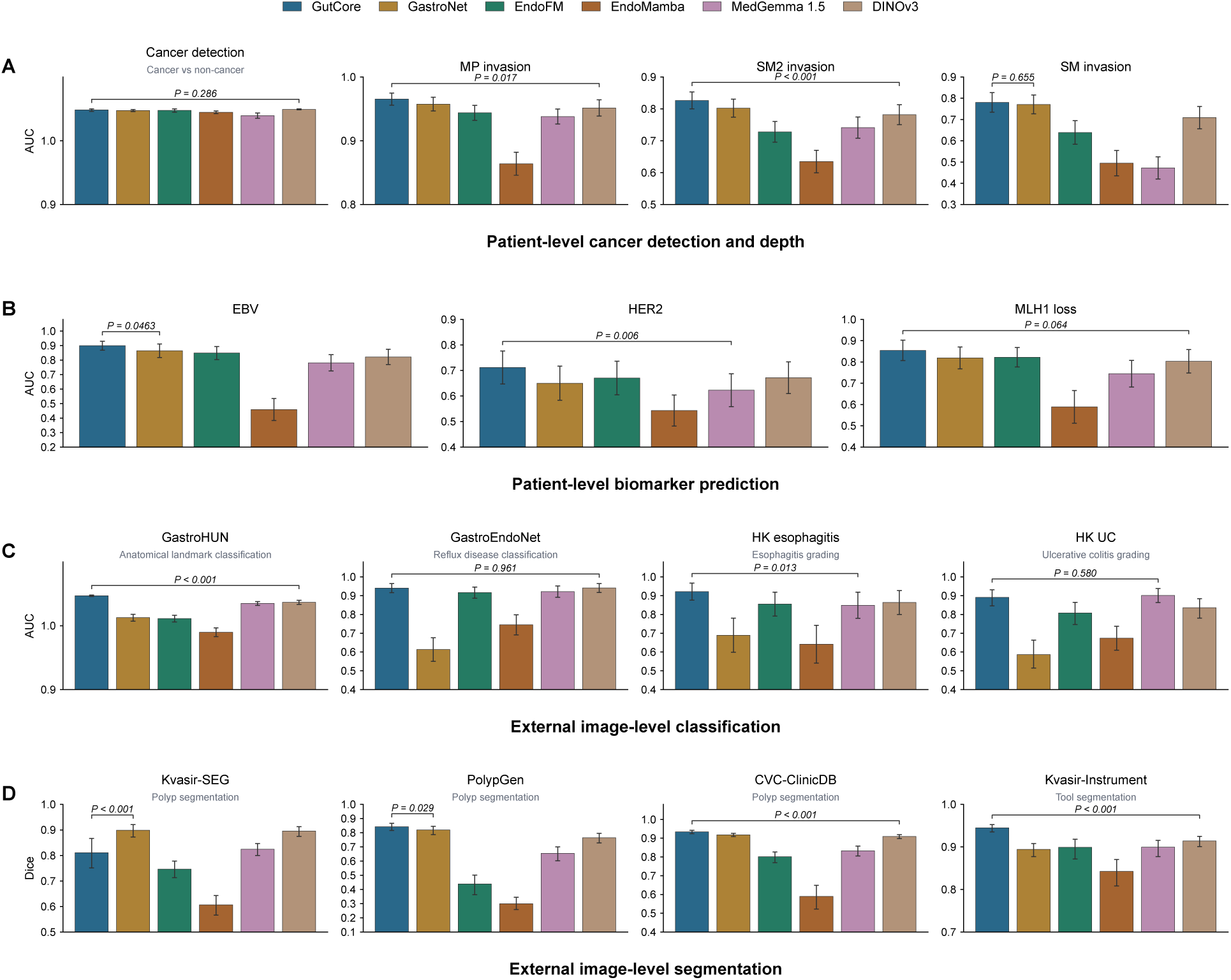
Task-specific performance after end-to-end fine-tuning. The same tasks and comparison models are shown as in Figure 2. Bars show held-out test-set performance, and error bars show 95% confidence intervals based on test images or patients. AUC is reported for classification and patient-level prediction tasks; Dice coefficient is reported for segmentation. Bracketed *P* values compare GutCore with the non-GutCore model that had the highest validation-set performance. Binary AUCs use paired DeLong tests; multiclass AUCs and Dice coefficients use two-sided paired bootstrap tests.

#### S4.2. Training-Time Image Subsampling

Patient-level endoscopy cases can contain many images, making repeated case-level training more computationally expensive than single-image classification. We therefore used random training-time image subsampling as a practical way to reduce computation. For each case-level training step, only a fixed maximum number of images was sampled from the available images in a case, while validation and test evaluation used all available case images. This design reduced the number of image features processed during training and randomized the images seen by the attention-based multiple-instance model, which can act as a regularization mechanism.

To examine whether this choice changed performance, we varied the maximum number of sampled training images per case across 4, 8, 12, 16, and 20 images for the four patient-level gastric cancer tasks using GutCore frozen-feature linear evaluation. Performance was similar across settings, and the 8-image setting reduced computation without a consistent loss of predictive performance. Accordingly, the main patient-level classification experiments used up to 8 sampled training images per case, with all available images retained at validation and test time.

**Figure S2:**
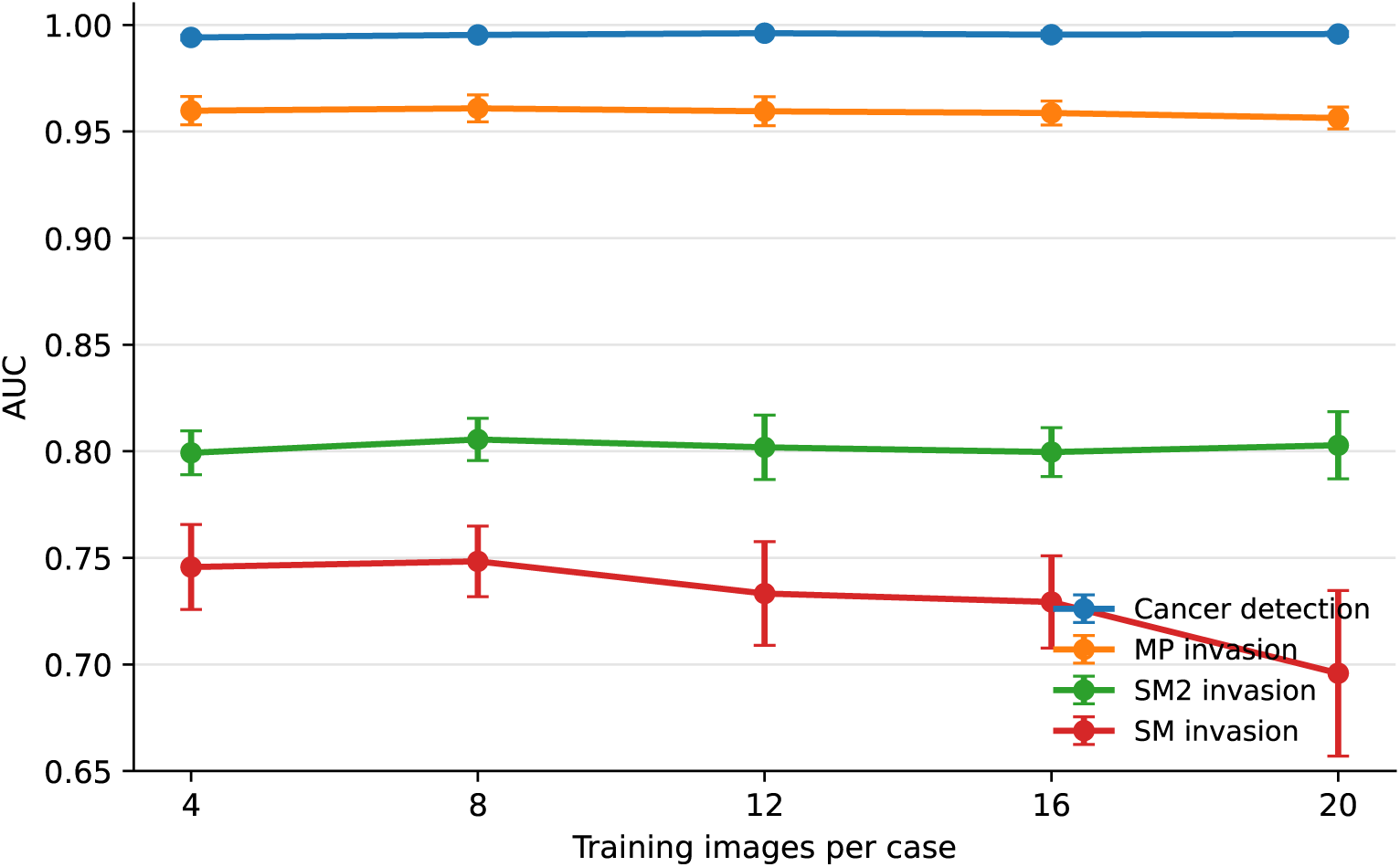
Training-time image subsampling for GutCore patient-level linear evaluation. The x-axis shows the maximum number of randomly sampled training images per case during training. Validation and test evaluation used all available images per case. Points and error bars show mean ± standard deviation of test AUC across five random train/validation/test splits.

